# Predicting Progression Free Survival after Systemic Therapy in Advanced Head and Neck Cancer: Bayesian regression and Model development

**DOI:** 10.1101/2021.09.22.21263942

**Authors:** Paul R Barber, Fabian Flores-Borja, Giovanna Alfano, Kenrick Ng, Gregory Weitsman, Luigi Dolcetti, Rami Mustapha, Felix Wong, Jose M Vicencio, Myria Galazi, James W Opzoomer, James N Arnold, Shahram Kordasti, Jana Doyle, Jon Greenberg, Magnus T Dillon, Kevin J Harrington, Martin D Forster, Anthony C C Coolen, Tony Ng

**Affiliations:** UCL Cancer Institute, Paul O’Gorman Building, University College London, London, UK; Comprehensive Cancer Centre, School of Cancer & Pharmaceutical Sciences, King’s College London, London, UK; Breast Cancer Now Research Unit, School of Cancer & Pharmaceutical Sciences, King’s College London, London, UK; Richard Dimbleby Laboratory of Cancer Research, School of Cancer & Pharmaceutical Sciences, King’s College London, London, UK; Tumor Immunology Group, School of Cancer & Pharmaceutical Sciences, King’s College London, London, UK; Systems Cancer Immunology, School of Cancer & Pharmaceutical Sciences, King’s College London, London, UK; Daichii Sankyo Incorporated, New Jersey, USA; The Institute of Cancer Research, London, UK; Institute for Mathematical and Molecular Biomedicine, King’s College London, UK; Saddle Point Science Ltd, London, UK

**Author notes:** To whom correspondence should be addressed: **Prof. Tony Ng**, (KCL). These authors contributed equally to this work (Paul R Barber, Fabian Flores-Borja, Giovanna Alfano, Kenrick Ng). Centre for Immunobiology and Regenerative Medicine, Barts & The London School of Medicine and Dentistry, Queen Mary University of London, London, UK.

## Abstract

**Background:** Advanced Head and Neck Squamous Cell Cancer (HNSCC) is associated with a poor prognosis, and biomarkers that predict response to treatment are highly desirable. The primary aim was to predict Progression Free Survival (PFS) with a multivariate risk prediction model.

**Methods:** Blood samples from 56 HNSCC patients were prospectively obtained within a Phase 2 clinical trial (NCT02633800), before and after the first treatment cycle of platinum-based chemotherapy, to identify biological covariates predictive of outcome. A total of 42 baseline covariates were derived pre-treatment, which were combined with 29 covariates after one cycle of treatment. These covariates were ranked and selected by Bayesian multivariate regression to form risk scores to predict PFS, producing “baseline” and “combined” risk prediction models respectively.

**Results:** The baseline model comprised of CD33+CD14+ monocytes, Double Negative B cells and age, in a weighted risk signature which predicted PFS with a concordance index (C-index) of 0.661. The combined model composed of baseline CD33+CD14+ monocytes, baseline Tregs, after-treatment changes in CD8 effector memory T cells, CD8 Central memory T cells and CD3 T Cells, along with the hypopharyngeal primary tumor subsite. This weighted risk signature exhibited an improved C-index of 0.757. There was concordance between levels of CD33+CD14+ myeloid cells in tumor tissue, as demonstrated by imaging mass cytometry, and peripheral blood in the same patients. This monocyte subpopulation also had univariate predictive value (log-rank p value = 0.03) but the C-index was inferior to the combined signature.

**Conclusions:** This immune-based combined multimodality signature, obtained through longitudinal peripheral blood monitoring, presents a novel means of predicting response early on during the treatment course.

**Funding:** Daiichi Sankyo Inc, Cancer Research UK, EU IMI2 IMMUCAN, UK Medical Research Council, European Research Council (335326), National Institute for Health Research and The Institute of Cancer Research.

## INTRODUCTION

Recurrent (R) or metastatic (M) Head and Neck Squamous Cell Carcinoma (HNSCC) is associated with a poor prognosis. Until the KEYNOTE-048 study was published in 2019(1), the standard-of-care, first line systemic treatment was the EXTREME regimen, consisting of a platinum-based chemotherapy regimen and cetuximab, an anti-EGFR monoclonal antibody(2). Even now, for patients with programmed death ligand 1 (PD-L1) negative tumors or those with contraindications to the use of anti-PD1 immunotherapy, the EXTREME regimen remains a first line standard-of-care.

While effective, this regimen is associated with toxicities. One of the key challenges for the treating physician is to identify the patients who would benefit from this treatment regimen. A predictive biomarker signature for patients with advanced HNSCC will help individualize discussions with patients regarding the risk-benefit balance of this treatment regimen and may guide patients who are likely to perform poorly towards alternative therapy regimens or clinical trials.

The absence of predictive biomarkers in this patient cohort represents a significant clinical unmet need. Until the development of PD-L1 as a biomarker for immunotherapy, efforts to generate biomarkers in HNSCC have focused on gene-expression profiles, which are dependent on the availability of tumor tissue and are only performed on pre-treatment samples(3, 4). Signatures based on a single biological modality and taken at a single timepoint may be insufficient to predict outcomes, as response to therapy relies on a dynamic interplay between cancer genomics, immune profile, tumor microenvironment, and clinicopathological characteristics of the patient receiving treatment(5, 6).

Efforts to develop a machine learning model to stratify survival risk by combining genetic and clinicopathological characteristics have revealed some success in advanced oral squamous cell carcinoma(7). We hypothesize that a multimodal analysis, taking into account both clinicopathological and laboratory-based biological covariates at different timepoints, would provide better predictive value.

We prospectively collected peripheral blood samples from a Phase 2 trial in R/M HNSCC (NTC02633800)(8), which utilized a modified EXTREME regimen as a backbone, and conducted a parallel exploratory analysis with the aim of generating a biomarker signature which would predict outcomes to treatment. We hypothesized that the detailed definition of a broad immune cell signature could contribute to the development of assays employing liquid biopsies to predict clinical outcomes. We also incorporated the analysis of two circulating microRNAs (miRNAs); miR-21-5p and miR-142-3p, which have previously demonstrated prognostic and predictive utility(9, 10). As the trial investigated the efficacy of an anti-ErbB3 antibody, patritumab, administered alongside an anti-EGFR antibody, we simultaneously analyzed EGFR-ErbB3 dimerization using Förster Resonance Energy Transfer (FRET) and included it in our analysis.

By extracting information from patient samples at baseline and after the first cycle of treatment within this trial, we aimed to develop a multimodal predictive signature for the EXTREME regimen based on a novel Bayesian multivariate model. This can serve as a non-invasive risk stratification for patients with R/M HNSCC using only peripheral blood, guiding the clinician towards the likelihood of success early during the treatment course.

## MATERIALS AND METHODS

### Study Design

The clinical study design of the Phase 2 study (NCT02633800) and its associated exploratory analysis are shown in Figure 1A. 87 patients were enrolled in the clinical trial. Peripheral blood samples were collected before initiation of treatment (C1) and immediately before the second cycle of treatment (C2). 31 patients were excluded due to incomplete paired biological datasets, leaving 56 patients for analysis. Amongst these patients, there was no difference in PFS as demonstrated by Kaplan-Meier survival curve analysis (Supplementary Figure 1) regardless of whether the patients received patritumab, which reflected the results published in the clinical trial. The baseline clinical characteristics of these 56 patients are shown in Supplementary Table 1.

**Figure 1:**
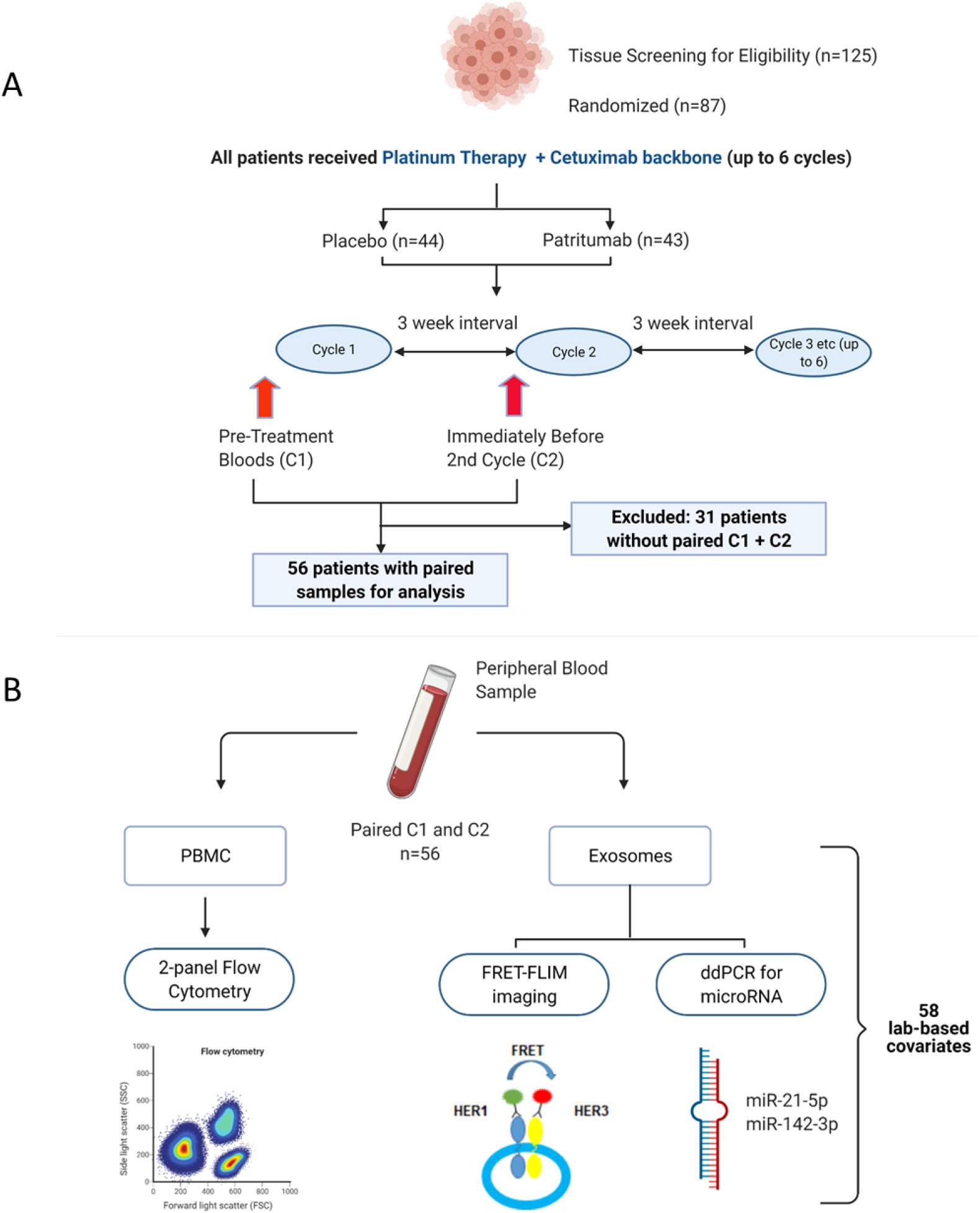
Peripheral blood samples from the clinical trial were prospectively analyzed using a multimodality platform. (A) Schematic of clinical trial design and timepoints at which peripheral blood was obtained. (B) Fifty-six (n=56) paired blood samples, obtained pre-treatment (C1) and after one cycle of treatment (C2) were subjected to Flow Cytometry, FRET-FLIM imaging and ddPCR analysis.

PBMC samples were analyzed using Flow Cytometry to generate unique immunological subpopulations. Exosomes were extracted from the serum and analyzed for EGFR-ErbB3 dimerization and miRNA-21-5p and miRNA-142-3p (Figure 1B). These analyses yielded a total of 29 unique biological covariates. Each covariate was obtained in pairs (C1 and C2), generating a total of 58 laboratory-based covariates for the multivariate analysis (Figure 1B). To mitigate individual baseline variations between patients, we evaluated changes between C1 and C2 (in the form of log-fold change, LFC, of the variable of interest) instead of absolute values of those parameters. A list of the laboratory-based and clinical covariates is provided in Supplementary Table 2.

The baseline clinical characteristics, as well as value of the laboratory-based covariates at baseline and after one cycle of treatment, did not significantly differ between the placebo and patritumab cohorts (Supplementary Table 3). Therefore, in this exploratory analysis, samples from both the control and investigational arms were analyzed together. The effect of adding the investigational product, patritumab on Progression-Free Survival (PFS) was evaluated by including it as an independent clinical covariate, denoted as ‘Drug’, in our multivariate analysis. Written informed consent was obtained. Approval was obtained from ethics committees (Research Ethics Committee reference: 15/LO/1670).

### Statistical Analysis

To examine whether the various features and distribution of survival indices indicated different prognostic outcomes, we built a model for predicting Progression-Free Survival (PFS). Covariates were ranked by importance and selected by Bayesian multivariate proportional hazards regression with backward elimination(11). We derived two models using separate datasets: firstly, a baseline predictive model containing a dataset of 42 baseline covariates (29 laboratory parameters at baseline, C1, and 13 clinical characteristics). The second, a combined predictive model, consists of 71 covariates, i.e. the 42 baseline covariates and a further 29 derived from the change in lab-based parameters between C1 and C2, measured by log fold change (LFC) of the variable of interest.

The relative efficiency of the predictive model was assessed by using C-index (a metric proposed by Harrell(12) to evaluate the accuracy of predictions made by an algorithm) and rank correlation of the signature-generated risk scores with survival time. The number of significant covariates in each prediction signature was determined with the aim of avoiding overfitting of the signature to the study data using the “batch regression” option of the Saddle Point Signature software (Saddle Point Science Ltd., London, UK), according to methods that were previously published(13, 14). This is particularly important with small number of patients where an independent test set is not possible. Systematic iterative covariate rejection and cross-validation (5000 iterations) allowed for the selection of an optimal covariate set to avoid overfitting though inclusion of too many covariates. The optimal set can be chosen in two ways, either based on the peak prediction performance of cross-validation, or the more stringent method that equally penalizes validation performance and overfitting (defined as the deviation between training and validation performance). All signatures presented were chosen using the more stringent criterion and data for all covariates is also presented for the purposes of identifying covariates that may be important but do not quite meet the criterion. The regression included covariates representing the missingness of data to account for the possibility that patient or sample selection/rejection (for any reason) is biased with respect to outcome and therefore could be informative. The missing data was imputed with the mean for that covariate. The importance and significance of covariates can be judged by their assigned beta value (β) in the proportional hazards model, and corresponding hazard ratio (HR) equal to e^2β^. A negative beta value reflects a lower risk of developing an event. The Signature software additionally judges the performance of similar randomized data, which most often has beta values around zero and within a critical range, such that any real covariate that has a beta value outside this critical range can be judged to be performing significantly better than randomized data. This adds additional confidence in the absence of an independent validation test set.

### Flow Cytometry

Frozen PBMC samples were thawed and stained with Fixable viability dye (Yellow Live/Dead^™^, Fisher Scientific) followed by two different panels of membrane markers. A panel for T cells included CD3, CD4, CD8, CD25, CD45RO, CD127, CCR7, and HLA-DR. A panel for B cells and monocytes included CD3, CD19, CD24, CD38, CD27, IgD, CD33, CD11b, CD14 and CD16 (full list of both antibody panels in Supplementary Table 4). These two panels allow definition of immune cell populations as described in Supplementary Figure 2. Patients’ samples and corresponding Fluorescence Minus One (FMO) Controls were acquired in a Fortessa II flow cytometer (BD, Berkshire, UK) and analyzed with FlowJo software (Tree Star). Populations were quantified as proportions of their respective parent population.

### Isolation of Serum exosomes

Exosomes were prepared using an optimized centrifugation method(15). Diluted serum was centrifuged at 300xg for 10 min to remove cell debris, 5000xg for 20 min to remove large vesicles and membrane fragments, and 12,200xg for 30 min to deplete microvesicles. This was followed by 100,000xg ultracentrifugation for 120 min at 4 °C to pellet exosomes with a TLA-55 rotor (Beckman Coulter). After a second 100,000xg ultracentrifugation for 60 minutes, the resulting pellets were washed and resuspended in PBS. Purified exosomal fractions were diluted and used for nanoparticle tracking analysis (NTA) using a Nanosight LM-14 system.

### RNA Extraction and miRNA Expression Analysis

RNA from cancer patients’ serum exosomes was extracted using the TRIzol™ Plus RNA Purification Kit (Thermo Fisher, UK) according to the manufacturer’ instructions. Quantification of gene expression in circulating exosomes was performed by ddPCR (Bio-Rad QX100 system). Normalization of the RNA, between cycle 1 and cycle 2 therapy of each patient, was performed using the expression levels of the housekeeping gene *18S* (Assay ID, Hs99999901_s1). For each sample, equal volume of RNA was used as template and cDNA synthesis performed using the SuperScript® VILO™ MasterMix (Thermo Fisher, UK) according to the manufacturer’ instructions. MicroRNAs were reverse-transcribed individually using the TaqMan™ MicroRNA Reverse Transcription Kit (Thermo Fisher, UK). For each sample, the normalized amount of RNA was reverse transcribed in a 15 μl reaction using the standard protocol and primers specific for each miRNA: miR-21-5p (assay ID, 000397), miR-142-3p (assay ID, 000464). Then 7.5 μl of cDNA was added to a 20 μl reaction containing 12.5 μl 2X ddPCR Supermix for Probes (Bio-Rad) and 1 μl 20X TaqMan miRNA PCR primer probe set; each reaction was carried out in duplicate. Thermo cycling conditions were as it follows: 95 °C for 10 min, then 50 cycles of 95 °C for 10 sec and 61 °C for 30 sec and a final inactivation step at 98 °C for 12 min. PCR products were analyzed using the QuantaSoft™ Software (Bio-Rad).

### ErbB3-EGFR Dimer Quantification in Exosomes

Exosomes were imaged on an ‘Open’ Fluorescence Lifetime Imaging Microscopy (FLIM) system (16). Analysis was performed with the TRI2 software (v2.7.8.9, CRUK/MRC Oxford Institute for Radiation Oncology, Oxford) as described previously (17, 18). Interfering effects of autofluorescence were minimized with a lifetime filtering algorithm and the FRET efficiency value for each patient calculated by: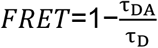, where t_D_ and t_DA_ are the average lifetime of Alexa Fluor546 in the matching donor (D) and donor-acceptor (DA) images.

### Imaging Mass Cytometry

FFPE histological slides were stained with a panel of metal conjugated antibodies (full list of antibodies listed in Supplementary Table 5).

In brief, antigen retrieval was performed on a Ventana Bench Mark Ultra with CC1 buffer (Roche, 950-224). Slides were blocked for 1 hour at RT in 5% BSA, 5mg/ml human IgG in PBS and stained overnight at 4°C in 4% BSA, PBS. DNA counterstain was performed with Iridium (Fluidigm, 201192B) 125nM in PBS for 30 minutes at room temperature.

Ablation and data acquisition were performed on a Fluidigm Hyperion located within our Biomedical Research Centre. Imaging analysis was performed using the RUNIMC R package: RandomForest for classification and regression, Raster and SF for image manipulation and segmentation (see https://www.biorxiv.org/content/10.1101/2021.09.14.460258v1, with code available here: https://github.com/luigidolcetti/RUNIMC).

## RESULTS

### The Model with Baseline Covariates Reveal Immune Subpopulations and Age Predict PFS

Bayesian multivariate proportional hazards regression was performed on the 42 covariates derived at baseline (C1) and PFS outcome. We utilized the stringent selection criteria based on a proportional hazards regression model to minimize overfitting based on the cross-validation performance (Figure 2A). This revealed two baseline immune subpopulations with a beta value which exceeded the critical beta-value threshold, ie CD14+CD16+CD33+CD11b+ monocytes (thereafter referred to as CD33+CD14+ monocytes according to previous nomenclature(19)) and double negative (CD27-IgD-) B cells (DN B cells), as well as one clinical covariate – age (Figure 2B). Missingness covariates were included in this analysis and did not affect the outcome of the signature.

**Figure 2.**
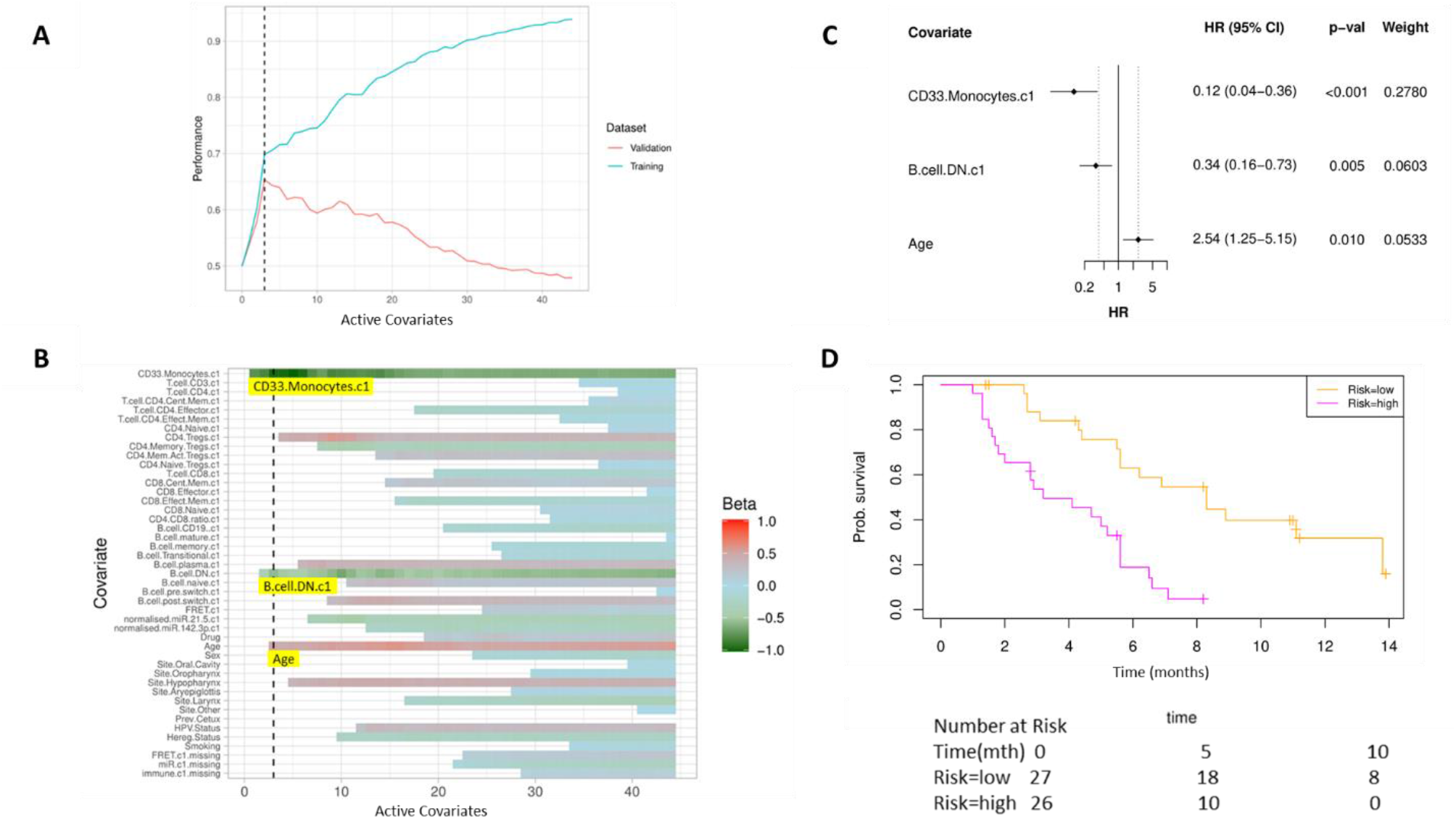
High baseline CD33+CD14+ monocytes and Double Negative B cells Predict Overall Survival. Covariates were ranked for importance and selected by a proportional hazards regression model with cross-validation and backward elimination. (A) Signature performance (defined as the classifier accuracy for predicting patient survival < or > median) as a function of the number of active covariates as covariates are eliminated based on cross validation performance. (B) Heatmap of covariate strength (beta value in the proportional hazards model) for the active covariates. Covariate elimination revealed three covariates in the optimal signature – CD33+CD14+ monocytes, Double Negative B Cells, and Age. (C) Forest plot of the three covariates within Progression Free Survival risk score with dotted line indicating the range, around 1, of typical random covariates. (D) Progression Free Survival Risk Signature Performance, low risk score (n=27) and high risk score (n=26). Log rank P-value = 6.0 e-5, with numbers at risk demonstrated under Kaplan-Meier curve. The multivariate analysis resulted in risk signatures that are linear combinations of weighted covariates. Their ability to predict outcome is demonstrated with data split by signature value.

Evaluation of the individual beta values reveal that baseline CD33+CD14+ monocytes and double negative B cells have a beta, β value of -1.05 and -0.53 respectively, and hence a higher baseline value of both populations is predictive of better PFS. Age, with a β value of 0.47, is associated with poorer PFS. The hazard ratios (HR) of the individual covariates are depicted in Figure 2C.

The risk scores generated from this signature were split at the median value to generate low-risk and high-risk cohorts (Figure 2D). The median overall survival of the low-risk and high-risk cohorts in this baseline predictive signature are 8.3 and 3.6 months respectively (log rank p-value = 6.0 e-5) with a rank correlation of -0.29. The C-index of the predictive signature based completely on baseline parameters is 0.661. The risk score equation is given in Supplementary Figure 4.

### Incorporating laboratory-based covariates after one cycle of treatment improves ability to predict PFS benefit

We subsequently evaluated if the incorporation of early laboratory-based changes into the signature improves its predictive ability. A separate predictive model incorporating 29 new covariates, ie changes in laboratory-based parameters between cycle 1 and cycle 2 was generated.

As before, we used a proportional hazards regression to determine a set of variables which predict PFS. A total of six covariates were identified – three immune subpopulations with negative beta values and hence are associated with better survival, ie baseline CD33+CD14+ monocytes, baseline CD4 memory regulatory T cells (HLA-DR^-^CD45RO^+^Tregs) and an increase in CD8 effector memory T cells (CD45RO^+^CCR7^-^). An increase in two subpopulations, CD8 Central Memory T Cells (CD45RO^+^CCR7^+^) and CD3 T cells, was associated with inferior PFS. The hypopharyngeal primary tumor site was also associated with a poorer PFS (Figure 3A and B).

**Figure 3.**
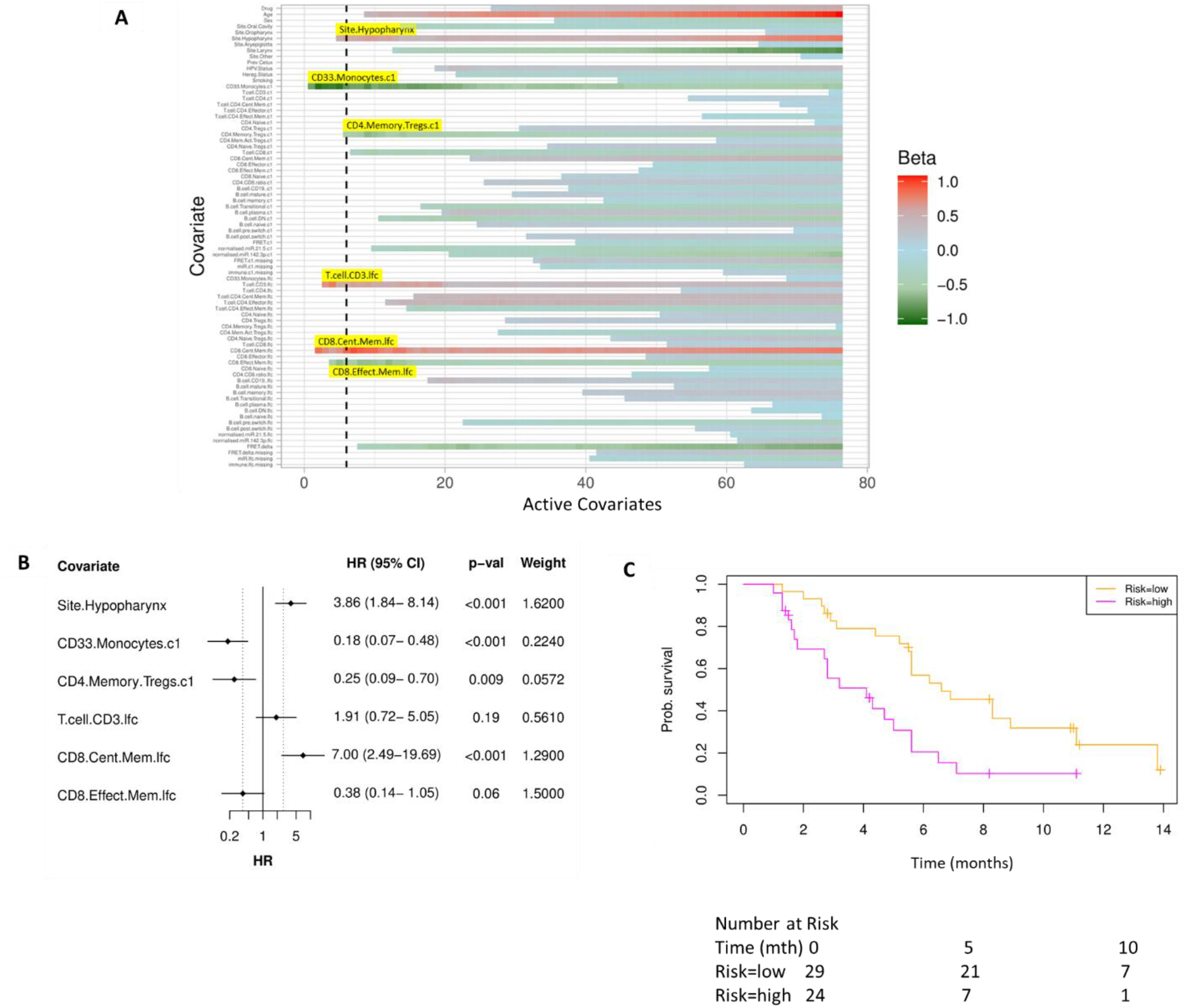
Model Incorporating Laboratory Changes After One Cycle of Treatment Exhibit Improved Predictive Value. (A) Heatmap of covariate strength (beta value in the proportional hazards model) for the active covariates. Proportional hazards regression revealed five immune subpopulations in the optimal model – Baseline CD33+CD14+ monocytes, Baseline CD4 Memory Regulatory T Cells, LFC of CD8 effector memory T cells, LFC of CD8 central memory T cells and LFC of CD3 T cells. The primary tumor site of hypopharynx also featured in the signature. A negative beta value is associated with lower risk score and hence better progression free survival. LFC = Log Fold Change (B) Forest plot of the three covariates within Progression Free Survival risk score (C) Progression Free Survival Risk Signature Performance, low risk score (n=29) and high risk score (n=24). Log rank P-value = 0.004, with numbers at risk demonstrated under Kaplan-Meier curve.

A multivariate analysis employing linear combinations of these six weighted covariates generated a risk signature. Their ability to predict outcome is demonstrated with data split by risk score, shown in Figure 3C. In this combined predictive signature, the median overall survival of the low-risk and high-risk cohorts are 6.8 and 3.6 months respectively (log rank p-value 0.004) with a rank correlation of -0.38 (Figure D). The C-index of the predictive signature, which incorporates baseline variables with changes in laboratory parameters after one cycle of treatment, is 0.757. Both values are greater in the combined signature when compared to the signature which only accounts for baseline values. The risk score equation is given in Supplementary Figure 4.

### EGFR-ErbB3 FRET may contribute to predictive signature

While the combined predictive signature comprised predominantly of immunological parameters, there is a suggestion that FRET difference may carry a degree of predictive value. In Figure 3A (4^th^ covariate from the bottom), the difference in EGFR-ErbB3 FRET (FRET.delta) was associated with a negative beta value which suggests a better PFS. However, the stringency that we have applied to optimal covariate selection means that this covariate fell marginally short of featuring in the eventual predictive signature. Nonetheless, this is the first time that this assay has been used within the context of a randomized controlled trial in exosomes and the suggested predictive value of the dimer warrants some discussion.

Figure 4A displays intensity images and donor lifetime map of exosomes labelled with anti-EGFR and anti-ErbB3 antibodies, along with an accompanying schematic (Figure 4B).

**Figure 4.**
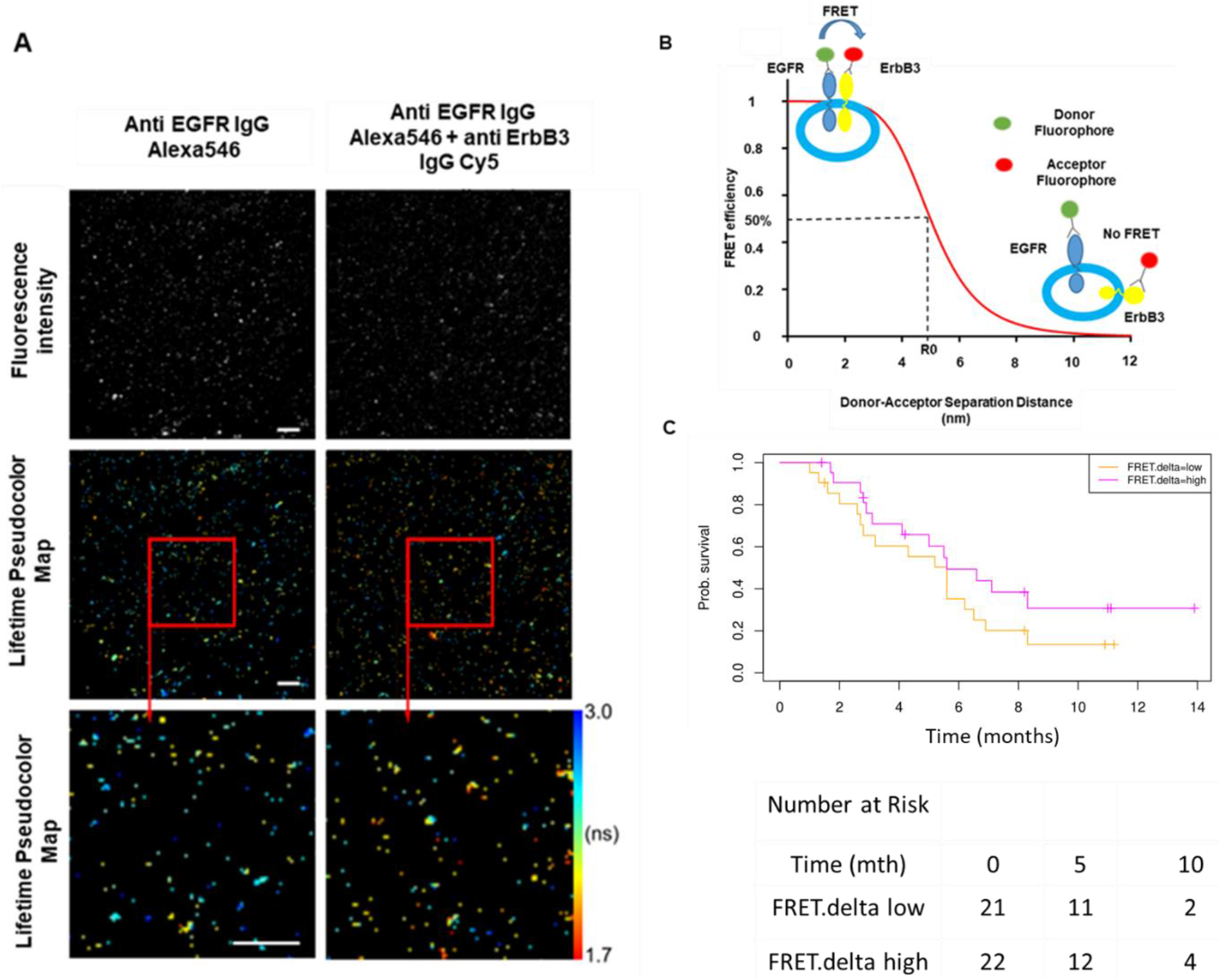
FRET/FLIM fluorescence assay of circulating exosomes extracted from patients. (A) Time-resolved fluorescence intensity images and donor lifetime map of exosomes labelled with Anti-EGFR-IgG-Alexa 546 and Anti-ErbB3-IgG-Cy5 extracellular antibodies (B) Schematic illustration of the fluorescent labelling geometry on exosomes and distance dependence of FRET efficiency (C) Progression Free Survival of subpopulations divided by median FRET difference, FRET.delta low (n=21) and FRET.delta high (n=22). Log rank P-value = 0.2, with numbers at risk demonstrated under Kaplan-Meier curve.

By dividing the patients with available FRET values by the median FRET.delta (n=43), there was a suggestion that patients with a high FRET.delta exhibited a better PFS than patients with a low FRET.delta. This difference was not statistically significant (p=0.2), and the predictive capacity of this univariate is limited (Rank Correlation = -0.132, C-index = 0.561, Figure 4C). Nonetheless, these results suggest a trend within a small patient cohort and can be explored in future prospective studies.

### Imaging Mass Cytometry of Tissue Reveals Correlation of CD33+CD14+ Myeloid Cell Subpopulation Between Tissue and Peripheral Blood

Having established that immune subsets in peripheral blood predict therapeutic response within a multivariate signature, we subsequently explored the relationship between the immune findings in peripheral blood with tumor infiltrating leukocytes (TILs). We obtained sufficient tissue from the biopsy at trial enrolment for in-depth profiling by imaging mass cytometry from four patients.

Standard FFPE samples from these four patients were processed as described previously, and the results clustered in an unsupervised fashion. The range of PFS for these patients was between 1.6-11.2 months (Figure 5A). Representative images of nuclear staining, overlaid with pixel-level classification, are shown for the four patients in Figure 5B. Heatmaps representing the distribution of cell phenotypes for each patient, as expressed by a two-phase classification conducted at pixel and cell level, are illustrated in Figure 5C. The list of cell populations characterized by imaging mass cytometry, alongside their detailed signature, is shown in Supplementary Table 6.

**Figure 5.**
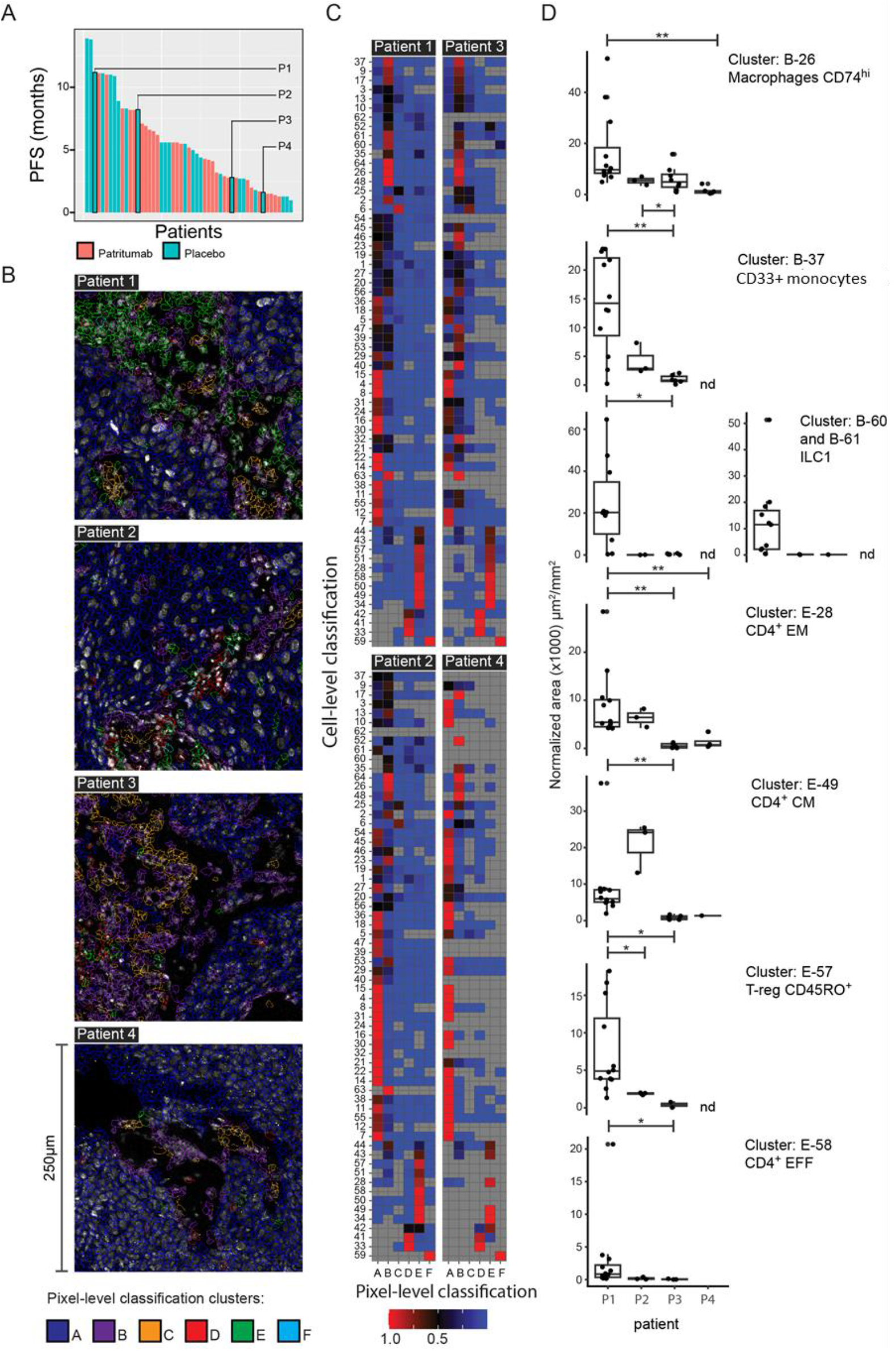
Image Mass Cytometry of Four Patient Samples Using a 29 Marker Panel Analysis. (A) Waterfall plot of Progression Free Survival showing the four patients used for imaging mass cytometry analysis (B) Representative image of the nuclear staining overlaid with pixel-level classification (C) Heatmaps representing the distribution of cell phenotypes for each patient, as expressed by a two-phase clustering conducted at pixel level (columns) and at cell level (rows). Data represent row-normalized areas. Red tiles, which represent hot spots of classification concordance, are further described in panel D (D) Differential analysis of the classification hotspots presented in panel C, highlighting cell populations which were significantly different between patients. Data are presented as sum of areas of positive cells normalized to the total area of the ROI and expressed as µm^2^ (x1000) per mm^2^. Statistics: *adj-p<0.05, **adj-p<0.01, ****adj-p<0.001, Pairwise Wilcoxon Rank Sum Tests with Benjamini correction, n≥3.

A total of 7 cell clusters identified on tissue mass cytometry were significantly different between the four patients. CD33+CD14+ monocytes (cluster B-37) which featured in both baseline and combined signatures, exhibited a diminishing trend of abundance across patients 1 to 4. Having observed this trend, we subsequently correlated the levels of this population in tissue with blood. The proportion of CD33+CD14+ monocytes in the blood was 17.4%, 5.39%, 2.0% and 2.47% for patients 1-4 respectively, suggesting a meaningful concordance between the levels of this subpopulation in the patient tissue and peripheral blood.

### CD33+CD14+ Monocyte Population have high HLA-DR expression and univariate predictive value

Due to the consistency with which the CD33+CD14+ monocyte population appeared across our study, we wanted to further characterize this population to determine its phenotype. During the process of drafting this manuscript, our lab was concurrently processing PBMCs from a cohort of patients at risk of developing lung cancer. Using a second flow cytometry staining panel which incorporates an alternative set of markers on these samples, we further characterized this monocyte subpopulation, affirming that these CD14+CD16+CD33+CD11b monocytes also express high levels of HLA-DR and CD11c (Supplementary Figure 3). This affirms that our population of interest closely resembles the previously described CD33+CD14+ monocytes(19).

MiRNA signatures have been implicated as a useful classifier for myeloid cell subsets(20). By correlating the miRNA changes in our study with this monocytic subpopulation, a significant correlation was identified between the log fold changes of miR-21-5p with the corresponding log fold changes of CD33+CD14+ monocytes (Pearson’s r=0.4343, p=0.02092, Figure 6A). We also investigated the potential of baseline CD33+CD14+ monocyte levels at predicting PFS. Figure 6B illustrates a Kaplan-Meier curve of PFS by median CD33+CD14+ monocyte level, generating a modest split which surpassed conventional statistical significance (log rank p-value = 0.03. However, the predictive capacity of CD33+CD14+ monocytes as a covariate was limited (C-index 0.593, rank-correlation 0.22). These values were inferior in predictive capacity compared to the baseline signature, which employed three covariates, and the combined predictive signature which employed six covariates (Figure 6C).

**Figure 6.**
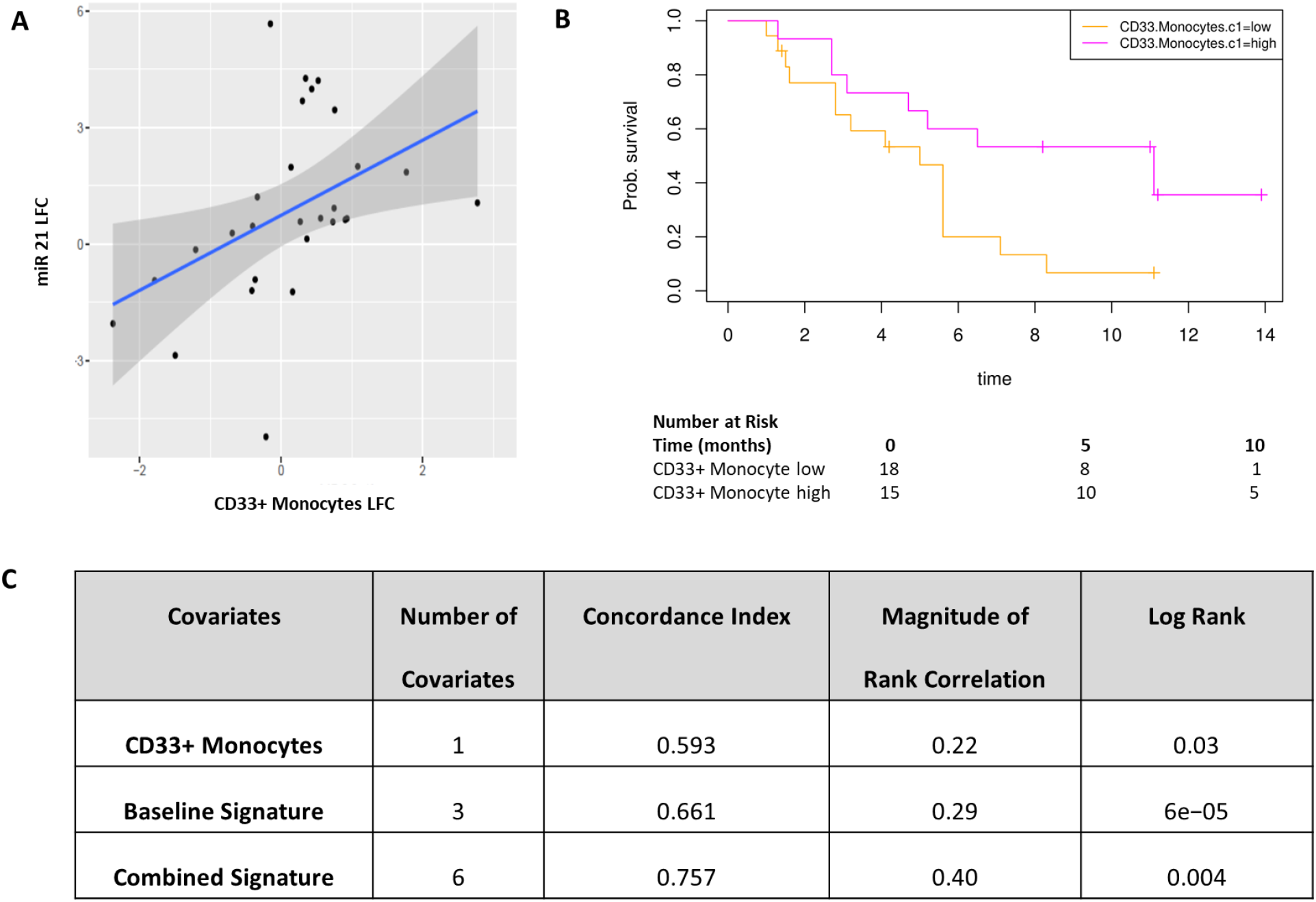
CD33+CD14+ Monocytes have some predictive value but predictive ability of signature is maximized with longitudinal sampling of peripheral blood. (A) (B) Correlation between miRNA 21 fold change and CD33+ monocyte fold change after one cycle of treatment. Correlation efficient of 0.4343, p value 0.02092. miR21: microRNA-21-5p, lfc: log-fold change (B) Kaplan-Meier Curve of PFS split by median CD33+CD14+ value, log rank p-value 0.03 (C) Table summarises C-index, rank correlation and log-rank p value based on type and number of covariates. There is an increasing C-index and rank correlation upon the strict selection of additional covariates into the predictive signature, with the strongest signature incorporating six covariates combining values from baseline and after one cycle of treatment.

## DISCUSSION

There is a clinical unmet need to identify predictive biomarkers for treatment in head and neck cancer. Gene expression profiling has revealed promising initial results in this domain but have been limited to HPV positive HNSCC, which inherently have better prognoses. Recent developments in the field of immunotherapy in HNSCC have focused on tissue-based biomarkers, such as PD-L1, but when used in isolation these have not been sufficiently predictive at identifying patients who would benefit(1).

While uni-modal biomarkers may offer some predictive value, the biology of HNSCC and likelihood of response to treatment is likely to be dictated by an interplay between tumor immunity, genomic signatures and a host of clinicopathological characteristics. There is an increased interest in peripheral blood biopsies in recent years, particularly in the context of peripheral blood mononuclear cells(21). The use of peripheral blood in deriving biomarkers mitigates a few limitations posed by tissue biopsies – particularly the accessibility and amount of tissue required. The ease of obtaining liquid biopsies also facilitates longitudinal monitoring of response to treatment.

To our knowledge, ours is the first piece of work integrating multiple biological covariates derived from peripheral blood to generate a signature which predicts treatment response. We also demonstrate the effectiveness of sequential monitoring of peripheral blood variables and the advantage of longitudinal monitoring at enhancing prediction of response, as shown by the combined predictive signature. By employing cross validation iterations to estimate training and validation errors, implementing advanced overfitting correlation protocols, using built-in corrections for informative data missingness, and probabilistic covariate removal, we were able to derive a robust optimal covariate set which correlates with PFS. This combination of analyses has been shown to produce robust signatures that do generalize to unseen data(13).

The biological components of the predicted model warrant discussion. The only clinical covariate to feature in the combined risk signature is the hypopharyngeal SCC sub-site, which was adversely correlated with PFS. This corroborates previous findings that the 5-year relative survival of patients with hypopharyngeal SCC is consistently the worst amongst different anatomical HNSCC sub-sites (22, 23). The propensity of hypopharyngeal tumors to present at the *de novo* advanced stage(24) and the density of submucosal lymphatics in this anatomical region translates into these patients inherently performing worse – lending support to the robust nature of our predictive signature. The notable absence of patritumab (denoted as ‘Drug’) in our predictive signatures is also consistent with the outcome of the Phase 2 Clinical Trial where the addition of this investigational medicinal product did not produce any benefit to PFS(8).

CD33+CD14+ monocytes demonstrated predictive capacity both as a univariate and as a prominent feature in both baseline and combined signatures. Monocytes are a heterogenous cell population, and phenotypic and functional characterization of monocyte subsets is a rapidly emerging field(25). Our identified population of interest, CD14+CD16+CD33+CD11b+CD11c+HLA-DR+ monocytes, resemble an intermediate monocyte phenotype. This subset remains one of the most poorly characterized monocytic subpopulations so far but have previously been linked to diverse immunological functions including antigen processing and presentation, angiogenesis, and monocyte activation(26). Interestingly, a significant correlation between the changes in miRNA-21-5p with changes in this monocytic subpopulation was detected, supporting previous suggestions that miRNA signatures can be a useful indicator of the functional state of myeloid cell subsets in cancer (20). The predictive capacity of this subpopulation warrants investigation and further characterization in future studies.

Beyond the interest in individual covariates, our study also reveals the potential of using liquid-based biological outcomes to predict outcome to therapy. It has been widely recognized that even the most utilized biomarkers, such as tumor PD-L1, have limited predictive value when used in isolation. Our study reveals that a targeted multimodality signature, obtained through longitudinal sampling of peripheral blood, is able to augment this predictive capacity and better identify patients who would benefit from a particular treatment regimen.

There are a few limitations to the study. The absence of overall survival (OS) within our current dataset represents one of the shortcomings of the study and it would have been interesting to assess whether the immune markers, particularly the CD33+CD14+ monocytic population, predict survival in the longer term. However, the accuracy of the predictive signature for OS would have been diluted by a variety of subsequent treatment regimens. Secondly, due to tissue scarcity, we only managed to obtain sufficient biopsy tissue from four patients for in-depth profiling by mass cytometry to investigate the correlation of the monocytic population between tissue and blood.

The present study shows that the combination of biomarkers established prospectively by liquid biopsies early in the treatment course offers potential for the provision of personalized treatments to patients (27). The post-stratification survival curves in our study demonstrate markedly different progression free survivals as a testament to this robust statistical model, and could represent an invaluable guide to clinicians during the initial stages of treatment.

## Data Availability

The data generated in this study and used for multivariate modelling are available from the UCL repository: https://doi.org/10.5522/04/16566207.v1

## NOTES

### Affiliations of authors

Breast Cancer Now Research Unit, School of Cancer & Pharmaceutical Sciences, King’s College London, London, UK (FF-B,TN); Richard Dimbleby Laboratory of Cancer Research, School of Cancer & Pharmaceutical Sciences, King’s College London, London, UK (GA,GW,FW,RM,TN); Comprehensive Cancer Centre, School of Cancer & Pharmaceutical Sciences, King’s College London, London, UK (PRB); UCL Cancer Institute, Paul O’Gorman Building, University College London, London, UK (PRB,JMV,MG,KN,JB,MDF,TN); Tumor Immunology Group, School of Cancer & Pharmaceutical Sciences, King’s College London, London, UK (JWO, JNA), Systems Cancer Immunology, School of Cancer & Pharmaceutical Sciences, King’s College London, London, UK (SK), Daiichi Sankyo Incorporated,New Jersey, USA (JD,JG); The Institute of Cancer Research/The Royal Marsden Hospital National Institute for Health Research Biomedical Research Centre, London, UK (KH, MTD); Institute for Mathematical and Molecular Biomedicine, King’s College London, UK (ACC), Saddle Point Science Ltd, London, UK (ACC), Centre for Immunobiology and Regenerative Medicine, Barts & The London School of Medicine and Dentistry, Queen Mary University of London, London, UK (Current Affiliation for FF-B) ; Barts Health NHS Trust, London, UK (Current Affiliation for MG).

## ETHICS AND PERMISSIONS

Written informed consent was obtained for all patients who participated in the Phase 2 clinical trial. Approval was obtained from ethics committees (Research Ethics Committee reference: 15/LO/1670). Approval to procure and process a separate cohort of blood samples from patients at risk of developing lung cancer was also obtained (IRAS ID: 261766).

## ACKNOWLEDGEMENTS

We thank the patients who participated in this Phase 2 trial and the staff members at the study sites who cared for them. We also thank Dr James Barrett for his assistance towards designing the statistical techniques applied in this study.

## PRIOR PRESENTATIONS

This study was presented at the 2018 American Society of Clinical Oncology (ASCO) Annual Meeting in Chicago, June 1-5, 2018.

## LIST OF SUPPLEMENTARY MATERIALS

**Supplementary Figure 1.**
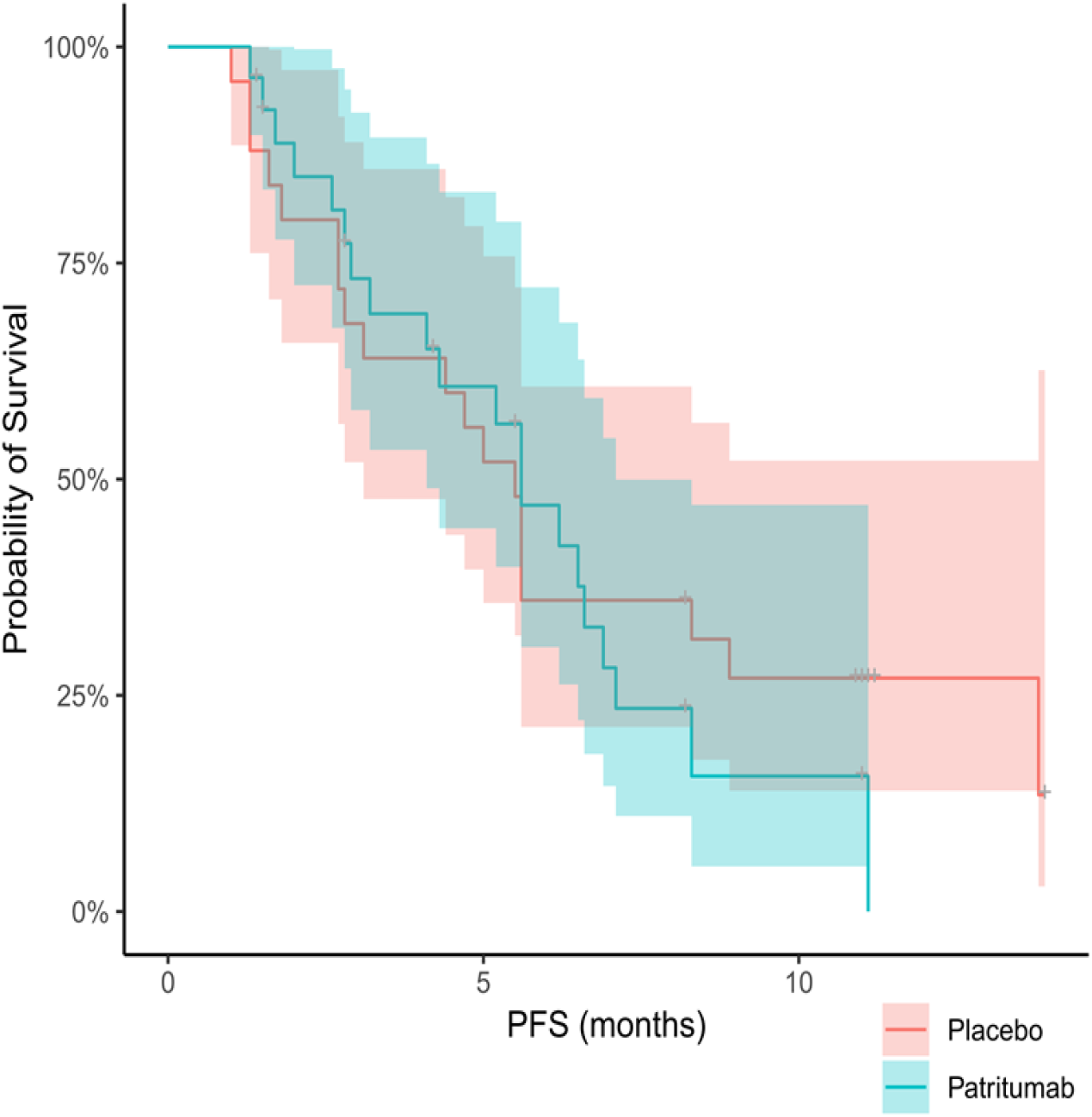
Kaplan Meier Curve of Progression-Free Survival in Study Cohort. Progression-Free Survival data was obtained from 53 patients, and stratified according to patients who received patritumab and the cohort which received placebo.

**Supplementary Table 1.** Demographic and Laboratory-Based Values of patients. Group of 56 patients with HNSCC was analyzed and their characteristics indicated as above.

|  | Overall |
| --- | --- |
| n | 56 |
| Age (mean (SD)) | 58.46 (8.78) |
| Drug = Placebo (%) | 27 (48.2) |
| Sex = M (%) | 46 (82.1) |
| Site (%) |  |
| Aryepiglottis | 1 ( 1.8) |
| Hypopharynx | 12 (21.4) |
| Larynx | 8 (14.3) |
| Oral Cavity | 20 (35.7) |
| Oropharynx | 10 (17.9) |
| Other | 5 ( 8.9) |
| Prev.Cetux = Yes (%) | 1 ( 1.8) |
| HPV.Status = Pos (%) | 8 (14.3) |
| Hereg.Status = Low (%) | 27 (48.2) |
| Smoking (%) |  |
| Current | 18 (32.1) |
| Ex | 34 (60.7) |
| Never | 3 ( 5.4) |
| Other | 1 ( 1.8) |
| Dendritic.CD33.c1 (mean (SD)) | 4.49 (4.46) |
| T.cell.CD3.c1 (mean (SD)) | 44.74 (20.79) |
| T.cell.CD4.c1 (mean (SD)) | 50.07 (18.43) |
| T.cell.CD4.Cent.Mem.c1 (mean (SD)) | 27.99 (9.58) |
| T.cell.CD4.Effector.c1 (mean (SD)) | 9.15 (18.07) |
| T.cell.CD4.Effect.Mem.c1 (mean (SD)) | 31.77 (17.04) |
| CD4.Naive.c1 (mean (SD)) | 30.07 (20.86) |
| CD4.Tregs.c1 (mean (SD)) | 8.97 (5.24) |
| CD4.Memory.Tregs.c1 (mean (SD)) | 62.15 (12.50) |
| CD4.Mem.Act.Tregs.c1 (mean (SD)) | 22.45 (12.25) |
| CD4.Naive.Tregs.c1 (mean (SD)) | 14.69 (14.08) |
| T.cell.CD8.c1 (mean (SD)) | 35.27 (12.98) |
| CD8.Cent.Mem.c1 (mean (SD)) | 9.34 (4.87) |
| CD8.Effector.c1 (mean (SD)) | 38.74 (15.99) |
| CD8.Effect.Mem.c1 (mean (SD)) | 33.38 (12.13) |
| CD8.Naive.c1 (mean (SD)) | 18.54 (13.92) |
| CD4.CD8.ratio.c1 (mean (SD)) | 1.85 (1.23) |
| B.cell.CD19..c1 (mean (SD)) | 17.15 (8.62) |
| B.cell.mature.c1 (mean (SD)) | 58.50 (13.45) |
| B.cell.memory.c1 (mean (SD)) | 21.18 (12.04) |
| B.cell.Transitional.c1 (mean (SD)) | 7.30 (8.30) |
| B.cell.plasma.c1 (mean (SD)) | 0.36 (0.62) |
| B.cell.DN.c1 (mean (SD)) | 13.84 (9.01) |
| B.cell.naive.c1 (mean (SD)) | 68.07 (14.55) |
| B.cell.pre.switch.c1 (mean (SD)) | 4.82 (5.52) |
| B.cell.post.switch.c1 (mean (SD)) | 13.26 (8.83) |
| FRET.c1 (mean (SD)) | 0.00 (0.05) |
| normalised.miR.21.5.c1 (mean (SD)) | 0.00 (0.01) |
| normalised.miR.142.3p.c1 (mean (SD)) | 0.00 (0.00) |
| FRET.c1.missing (mean (SD)) | 0.14 (0.35) |
| miR.c1.missing (mean (SD)) | 0.11 (0.31) |
| immune.c1.missing (mean (SD)) | 0.38 (0.49) |
| Dendritic.CD33.lfc (mean (SD)) | 0.31 (1.29) |
| T.cell.CD3.lfc (mean (SD)) | 0.28 (0.57) |
| T.cell.CD4.lfc (mean (SD)) | 0.12 (0.63) |
| T.cell.CD4.Cent.Mem.lfc (mean (SD)) | 0.01 (0.62) |
| T.cell.CD4.Effector.lfc (mean (SD)) | -0.37 (1.00) |
| T.cell.CD4.Effect.Mem.lfc (mean (SD)) | 0.01 (0.84) |
| CD4.Naive.lfc (mean (SD)) | 0.08 (0.98) |
| CD4.Tregs.lfc (mean (SD)) | -0.21 (0.72) |
| CD4.Memory.Tregs.lfc (mean (SD)) | -0.02 (0.30) |
| CD4.Mem.Act.Tregs.lfc (mean (SD)) | 0.19 (0.77) |
| CD4.Naive.Tregs.lfc (mean (SD)) | -0.10 (0.97) |
| T.cell.CD8.lfc (mean (SD)) | 0.11 (0.41) |
| CD8.Cent.Mem.lfc (mean (SD)) | 0.12 (0.74) |
| CD8.Effector.lfc (mean (SD)) | -0.17 (0.62) |
| CD8.Effect.Mem.lfc (mean (SD)) | 0.03 (0.33) |
| CD8.Naive.lfc (mean (SD)) | -0.15 (0.85) |
| CD4.CD8.ratio.lfc (mean (SD)) | -0.12 (0.79) |
| B.cell.CD19..lfc (mean (SD)) | -0.07 (0.89) |
| B.cell.mature.lfc (mean (SD)) | 0.03 (0.45) |
| B.cell.memory.lfc (mean (SD)) | 0.10 (0.45) |
| B.cell.Transitional.lfc (mean (SD)) | -0.26 (0.77) |
| B.cell.plasma.lfc (mean (SD)) | 0.12 (2.03) |
| B.cell.DN.lfc (mean (SD)) | -0.18 (0.61) |
| B.cell.naive.lfc (mean (SD)) | -0.01 (0.21) |
| B.cell.pre.switch.lfc (mean (SD)) | 0.14 (0.66) |
| B.cell.post.switch.lfc (mean (SD)) | 0.01 (0.62) |
| normalised.miR.21.5.lfc (mean (SD)) | 0.25 (2.12) |
| normalised.miR.142.3p.lfc (mean (SD)) | 0.13 (1.98) |
| FRET.delta (mean (SD)) | 0.00 (0.05) |
| FRET.delta.missing (mean (SD)) | 0.18 (0.39) |
| miR.lfc.missing (mean (SD)) | 0.11 (0.31) |
| immune.lfc.missing (mean (SD)) | 0.46 (0.50) |
| BOR.RECIST (mean (SD)) | 2.74 (0.68) |
| event.PFS (mean (SD)) | 0.74 (0.45) |
| t.PFS (mean (SD)) | 5.48 (3.48) |

**Supplementary Table 2:**
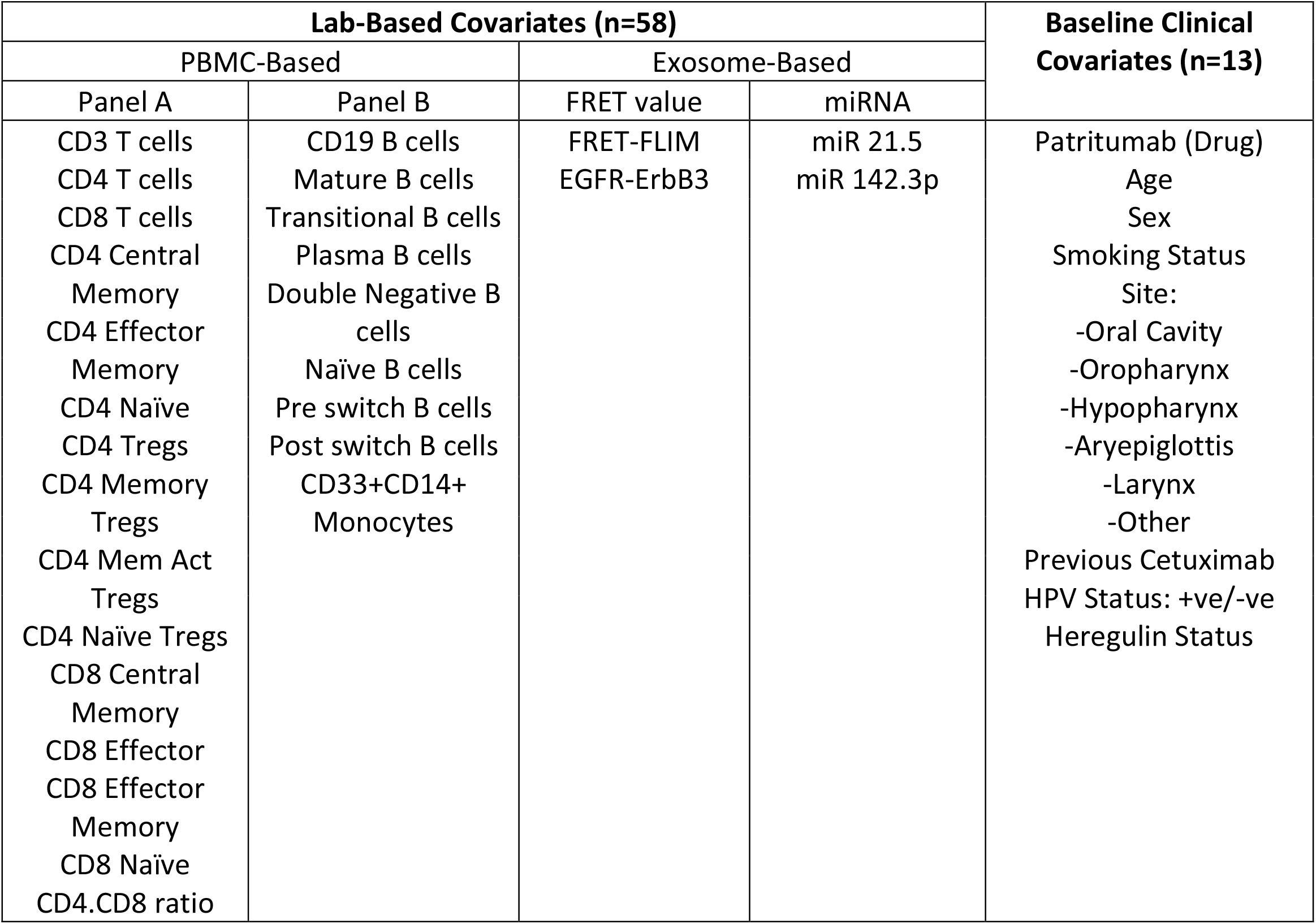
List of covariates entered into Bayesian multivariate analysis. 29 laboratory-based covariates were used (obtained at baseline and after one cycle of treatment), and combined with 13 baseline clinical covariates.

| Lab-Based Covariates (n=58) |  |  |  | Baseline Clinical Covariates (n=13) |
| --- | --- | --- | --- | --- |
| PBMC-Based |  | Exosome-Based |  |  |
| Panel A | Panel B | FRET value | miRNA |  |
| CD3 T cells<br>CD4 T cells<br>CD8 T cells<br>CD4 Central Memory<br>CD4 Effector Memory<br>CD4 Naïve<br>CD4 Tregs<br>CD4 Memory Tregs<br>CD4 Mem Act Tregs<br>CD4 Naïve Tregs<br>CD8 Central Memory<br>CD8 Effector<br>CD8 Effector Memory<br>CD8 Naïve<br>CD4.CD8 ratio | CD19 B cells<br>Mature B cells<br>Transitional B cells<br>Plasma B cells<br>Double Negative B cells<br>Naïve B cells<br>Pre switch B cells<br>Post switch B cells<br>CD33+CD14+ Monocytes | FRET-FLIM<br>EGFR-ErbB3 | miR 21.5<br>miR 142.3p | Patritumab (Drug)<br>Age<br>Sex<br>Smoking Status<br>Site:<br>-Oral Cavity<br>-Oropharynx<br>-Hypopharynx<br>-Aryepiglottis<br>-Larynx<br>-Other<br>Previous Cetuximab<br>HPV Status: +ve/-ve<br>Heregulin Status |

**Supplementary Table 3.**
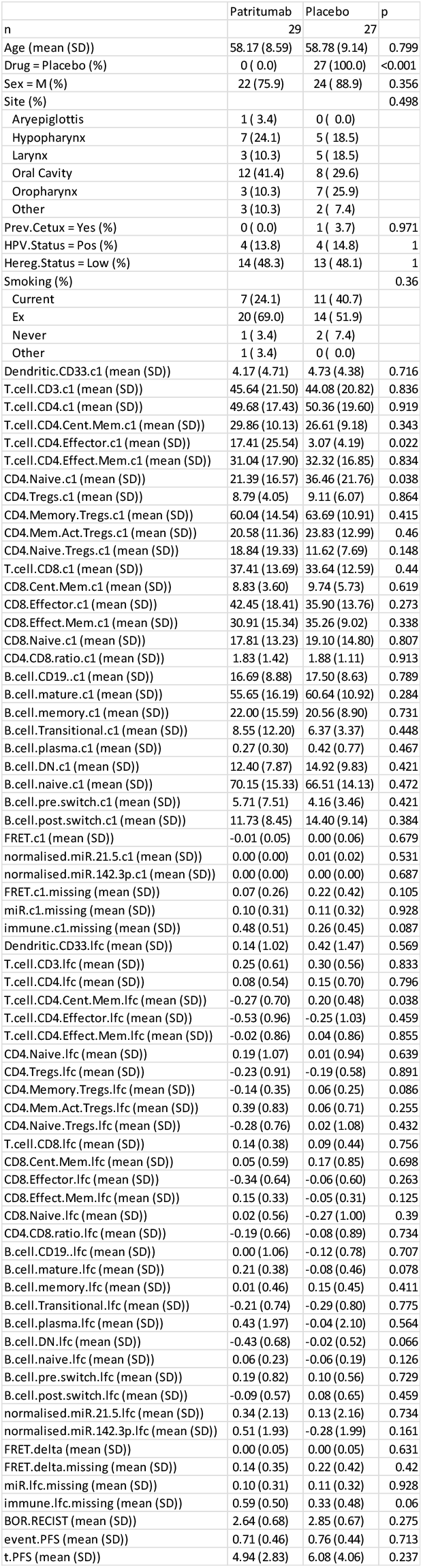
Demographic and Laboratory-Based Values of patients separated by arm of treatment on trial (placebo vs patritumab)

**Supplementary Figure 2.**
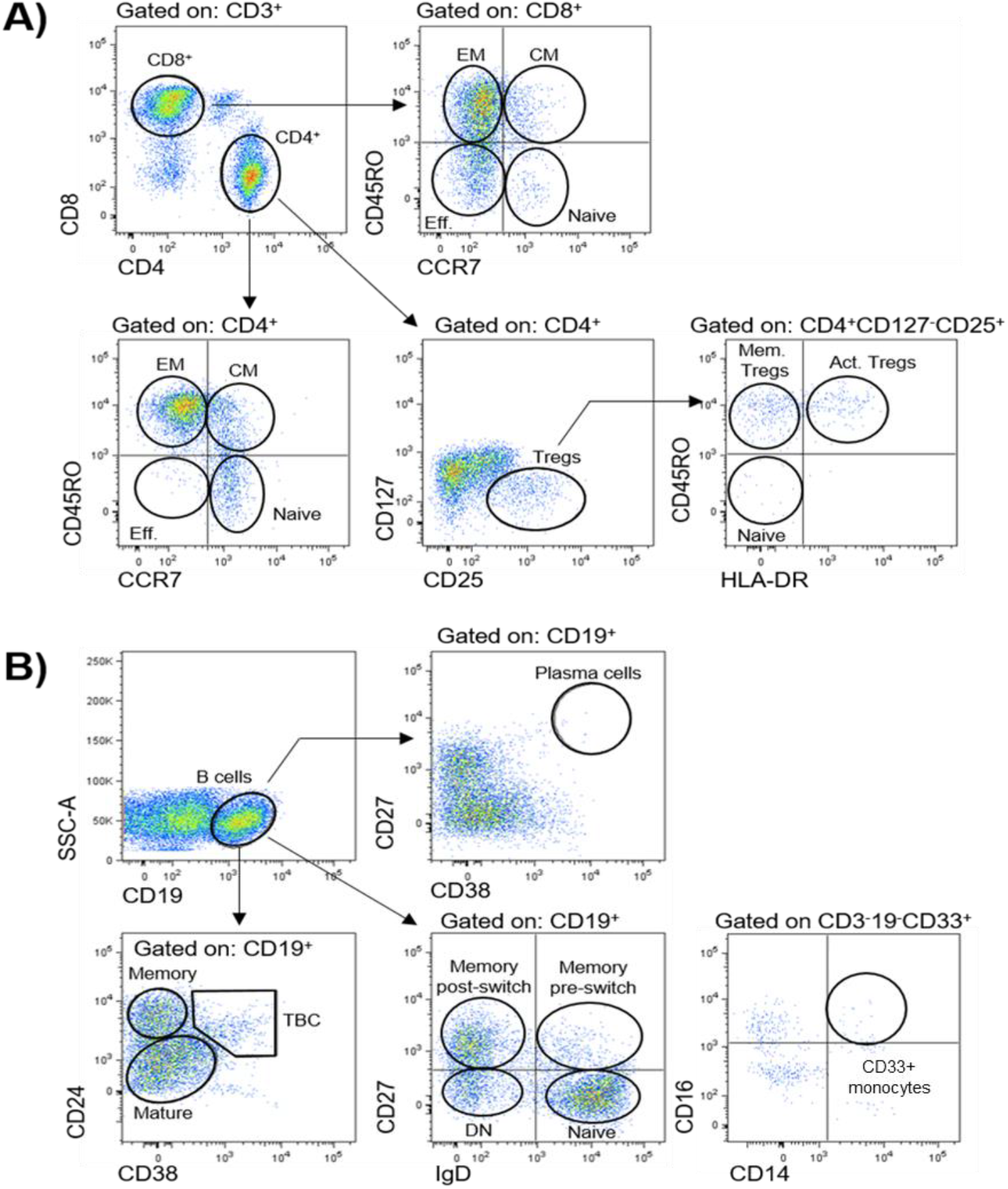
Gating strategies for definition of peripheral blood immune populations. Peripheral immune cell populations analysed in this study are indicated in corresponding gates **A)** Subpopulations of CD3^+^CD4^+^ and CD3^+^CD8^+^ T cells were further identified by the use of markers CCR7 and CD45RO as effector (Eff., CD45RO^-^CCR7^-^), naïve (CD45RO^-^ CCR7^+^), central memory (CM, CD45RO^+^CCR7^+^) and effector memory (EM, CD45RO^+^CCR7^-^). The activation status of CD4^+^ regulatory T cells (Tregs, CD127^-/lo^ CD25^hi^) was defined by markers CD45RO and HLA-DR as indicated: naïve Tregs (HLA-DR^-^CD45RO^-^), activated Tregs (Act. Tregs, HLA-DR^+^CD45RO^+^), and memory Tregs (Mem. Tregs, HLA-DR^-^CD45RO^+^). **B)** CD19^+^ B cell subpopulations were identified with markers CD24, CD38 and CD27: plasma cells (CD38+CD27^+^); mature (CD24^lo^CD38^lo^), memory (CD24^hi^CD38^-^), transitional (TBC, CD24^hi^CD38^hi^); or a combination of IgD and CD27:naïve (IgD^+^CD27^-^), memory pre-switch (IgD^+^CD27^+^), memory post-switch (IgD^-^CD27^+^), or double-negative B cells (DN, IgD^-^ CD27^-^). CD33+ CD14+ Monocytes were defined as CD3^-^ CD19^-^ CD33^+^ CD16^+^ CD14^+^.

**Supplementary Table 4.** List of antibodies used in T cell panel and B cell-monocyte panel for analyses of immune cell populations by flow cytometry. Antibodies were purchased from BD Biosciences and Biolegend as indicated.

| T cell Panel |  |  | B cell-Monocyte Panel |  |  |
| --- | --- | --- | --- | --- | --- |
| Target | Clone | Company | Target | Clone | Company |
| CD3 | UCHT1 | BD Biosciences | CD3 | UCHT1 | BD Biosciences |
| CD4 | SK3 | BD Biosciences | CD11b | ICRF44 | Biolegend |
| CD8 | RPA-T8 | BD Biosciences | CD14 | HCD14 | Biolegend |
| CD25 | M-A251 | BD Biosciences | CD16 | 3G8 | BD Biosciences |
| CD45RO | UCHL1 | BD Biosciences | CD19 | HIB19 | Biolegend |
| CD127 | HIL-7R-M21 | BD Biosciences | CD24 | ML5 | Biolegend |
| CCR7 | GO43H7 | Biolegend | CD27 | LI28 | Biolegend |
| HLA-DR | G46-6 | BD Biosciences | CD33 | P67.6 | Biolegend |
|  |  |  | CD38 | HB7 | Biolegend |
|  |  |  | IgD | IA6.2 | Biolegend |

**Supplementary Table 5.** List of antibodies used in mass cytometry (CyToF) analyses for definition of immune cell subpopulations in tissue.

| Channel | Target | CLONE | Company | Code | Custom Conjugation |
| --- | --- | --- | --- | --- | --- |
| 142Nd | SMA | 1A4 | Fluidigm | 3141017D | No |
| 144Nd | CD14 | EPR3653 | Fluidigm | 3144025D | No |
| 149Sm | CD11b | EPR1344 | Fluidigm | 3149028D | No |
| 150Nd | CD15 | W6D3 | Fluidigm | 3149026D | No |
| 151Eu | IgD | IgD26 | NovusBio | NBP2-50437 | Yes |
| 153Eu | CD24 | ALB9 | abcam | ab31622 | Yes |
| 154Sm | CD11c | EP1347Y | abcam | ab216655 | Yes |
| 160Gd | CD68 | KP1 | Fluidigm | 3159035D | No |
| 164Dy | ARGINASE-1 | D4E3M | Fluidigm | 3164027D | No |
| 165Ho | CD45RA | HI100 | BioLegend | 304102 | Yes |
| 166Er | CD74 | LN2 | Fluidigm | 3166025D | No |
| 175Lu | CD25 | EPR6452 | Fluidigm | 3175036D | No |
| 141Pr | CD38 | EPR4106 | Fluidigm | 3141018D | No |
| 143Nd | VIMENTIN | RV202 | Fluidigm | 3143029D | No |
| 145Nd | CD33 | POLY | Fluidigm | 3145017D | No |
| 146Nd | CD16 | EPR16784 | Fluidigm | 3146020D | No |
| 148Nd | Pan-Keratin | C11 | Fluidigm | 3148020D | No |
| 152Sm | CD45 | CD45-2B11 | Fluidigm | 3152016D | No |
| 155Gd | FOXP3 | 236A/E7 | Fluidigm | 3155016D | No |
| 156Gd | CD4 | EPR6855 | Fluidigm | 3156033D | No |
| 158Gd | E-CAD | 24 E10 | Fluidigm | 3158029D | No |
| 161Dy | CD20 | H1 | Fluidigm | 3161029D | No |
| 162Dy | CD8a | D8A8Y | Fluidigm | 3162035D | No |
| 168Er | CD127 | EPR2955(2) | Fluidigm | 3168026D | No |
| 169Tm | COLLAGEN-I | POLY | Fluidigm | 3169023D | No |
| 170Er | CD3 | POLY | Fluidigm | 3170019D | No |
| 171Yb | CD27 | EPR8569 | Fluidigm | 3171024D | No |
| 173Yb | CD45RO | UCHL1 | Fluidigm | 3173016D | No |

**Supplementary Table 6.** List of populations in imaging mass cytometry of tissue and corresponding signature. ILC = Innate Lymphoid Cells, T-reg = Regulatory T cells, SMA = Smooth Muscle Actin, MO-MDSC = Monocytic Myeloid Derived Suppressor Cells, G-MDSC = Granulocytic Myeloid Derived Suppressor Cells

| Pixel-level | Cell-level | Signature | Populations |
| --- | --- | --- | --- |
| A | 0 | e-Cadherin<sup>+</sup> / Pan-Keratinin<sup>+/−</sup> / Vimentin<sup>−</sup> | Epithelium/ tumor |
| B | 26 | CD11b<sup>int</sup> / CD33<sup>neg</sup> / CD14<sup>int</sup> / CD15<sup>neg</sup> / CD68<sup>hi</sup> / CD11c<sup>int</sup> / CD74<sup>hi</sup> | Macrophages CD74<sup>hi</sup> |
|  | 37 | CD11b<sup>int</sup> / CD33<sup>pos</sup> / CD14<sup>hi</sup> / CD15<sup>neg</sup> / CD68<sup>int</sup> / CD11c<sup>int</sup> / CD74<sup>int</sup> | CD33+ CD14+ Monocytes |
|  | 48 | CD11b<sup>int</sup> / CD33<sup>neg</sup> / CD14<sup>hi</sup> / CD15<sup>neg</sup> / CD68<sup>hi</sup> / CD11c<sup>int</sup> / CD74<sup>int</sup> | Mono/ Macro |
|  | 60 | CD11b<sup>int</sup> / CD33<sup>pos</sup> / CD14<sup>hi</sup> / CD15<sup>neg</sup> / CD68<sup>neg</sup> / CD11c<sup>neg</sup> / CD74<sup>int</sup> | MO-MDSC |
|  | 61 | CD11b<sup>int</sup> / CD33<sup>neg</sup> / CD14<sup>int</sup> / CD15<sup>neg</sup> / CD68<sup>neg</sup> / CD11c<sup>neg</sup> / CD74<sup>int</sup> | ILC-1 |
|  | 64 | CD11b<sup>hi</sup> / CD33<sup>neg</sup> / CD14<sup>int</sup> / CD15<sup>hi</sup> / CD68<sup>int</sup> / CD11c<sup>neg</sup> / CD74<sup>neg</sup> | G-MDSC |
|  | 9 | CD11b<sup>int</sup> / CD33<sup>pos</sup> / CD14<sup>int</sup> / CD15<sup>neg</sup> / CD68<sup>neg</sup> / CD11c<sup>neg</sup> / CD74<sup>neg</sup> | Monocyte |
|  | 0 | --- | Myeloid |
| C | 0 | SMA<sup>+</sup> | Fibroblast |
| D | 33 | CD3<sup>hi</sup> / CD8<sup>hi</sup> / CD27<sup>neg</sup> / CD45RA<sup>neg</sup> / CD45RO<sup>int</sup> / CD127<sup>neg</sup> | CD8<sup>+</sup> Effector |
|  | 41 | CD3<sup>hi</sup> / CD8<sup>hi</sup> / CD27<sup>int</sup> / CD45RA<sup>hi</sup> / CD45RO<sup>int</sup> | CD8<sup>+</sup> Central memory |
|  | 42 | CD3<sup>hi</sup> / CD8<sup>hi</sup> / CD27<sup>neg</sup> / CD45RA<sup>neg</sup> / CD45RO<sup>hi</sup> | CD8<sup>+</sup> Effector memory |
|  | 0 | --- | CD8<sup>+</sup> T-Cells |
| E | 28 | CD3<sup>low</sup> / CD4<sup>hi</sup> / CD27<sup>neg</sup> / CD45RA<sup>neg</sup> / CD45RO<sup>int</sup> / foxP3<sup>neg</sup> / CD25<sup>neg</sup> / CD127<sup>int</sup> / CD74<sup>hi</sup> | CD4<sup>+</sup> Effector memory |
|  | 34 | CD3<sup>hi</sup> / CD4<sup>hi</sup> / CD27<sup>neg</sup> / CD45RA<sup>neg</sup> / CD45RO<sup>hi</sup> / foxP3<sup>neg</sup> / CD25<sup>neg</sup> / CD127<sup>int</sup> / CD74<sup>int</sup> | CD4<sup>+</sup> Effector memory |
|  | 49 | CD3<sup>hi</sup> / CD4<sup>hi</sup> / CD27<sup>neg</sup> / CD45RA<sup>neg</sup> / CD45RO<sup>int</sup> / foxP3<sup>neg</sup> / CD25<sup>neg</sup> / CD127<sup>low</sup> / CD74<sup>int</sup> | CD4<sup>+</sup> Central memory |
|  | 50 | CD3<sup>hi</sup> / CD4<sup>hi</sup> / CD27<sup>int</sup> / CD45RA<sup>neg</sup> / CD45RO<sup>int</sup> / foxP3<sup>int</sup> / CD25<sup>low</sup> / CD127<sup>low</sup> / CD74<sup>int</sup> | CD4<sup>+</sup> activated |
|  | 51 | CD3<sup>hi</sup> / CD4<sup>hi</sup> / CD27<sup>hi</sup> / CD45RA<sup>hi</sup> / CD45RO<sup>int</sup> / foxP3<sup>neg</sup> / CD25<sup>low</sup> / CD127<sup>int</sup> / CD74<sup>int</sup> | CD4<sup>+</sup> Naïve |
|  | 57 | CD3<sup>hi</sup> / CD4<sup>hi</sup> / CD27<sup>int</sup> / CD45RA<sup>neg</sup> / CD45RO<sup>int</sup> / foxP3<sup>hi</sup> / CD25<sup>int</sup> / CD127<sup>low</sup> / CD74<sup>int</sup> | T-reg CD45RO+ |
|  | 58 | CD3<sup>hi</sup> / CD4<sup>hi</sup> / CD27<sup>int</sup> / CD45RA<sup>int</sup> / CD45RO<sup>int</sup> / foxP3<sup>neg</sup> / CD25<sup>low</sup> / CD127<sup>int</sup> / CD74<sup>int</sup> | CD4<sup>+</sup> Effector |
|  | 0 | --- | CD4<sup>+</sup> T-Cells |
| F | 59 | CD3<sup>neg</sup> / CD20<sup>hi</sup> | B-Cell |
|  | 0 | --- | B-Cell |

**Supplementary Figure 3.**
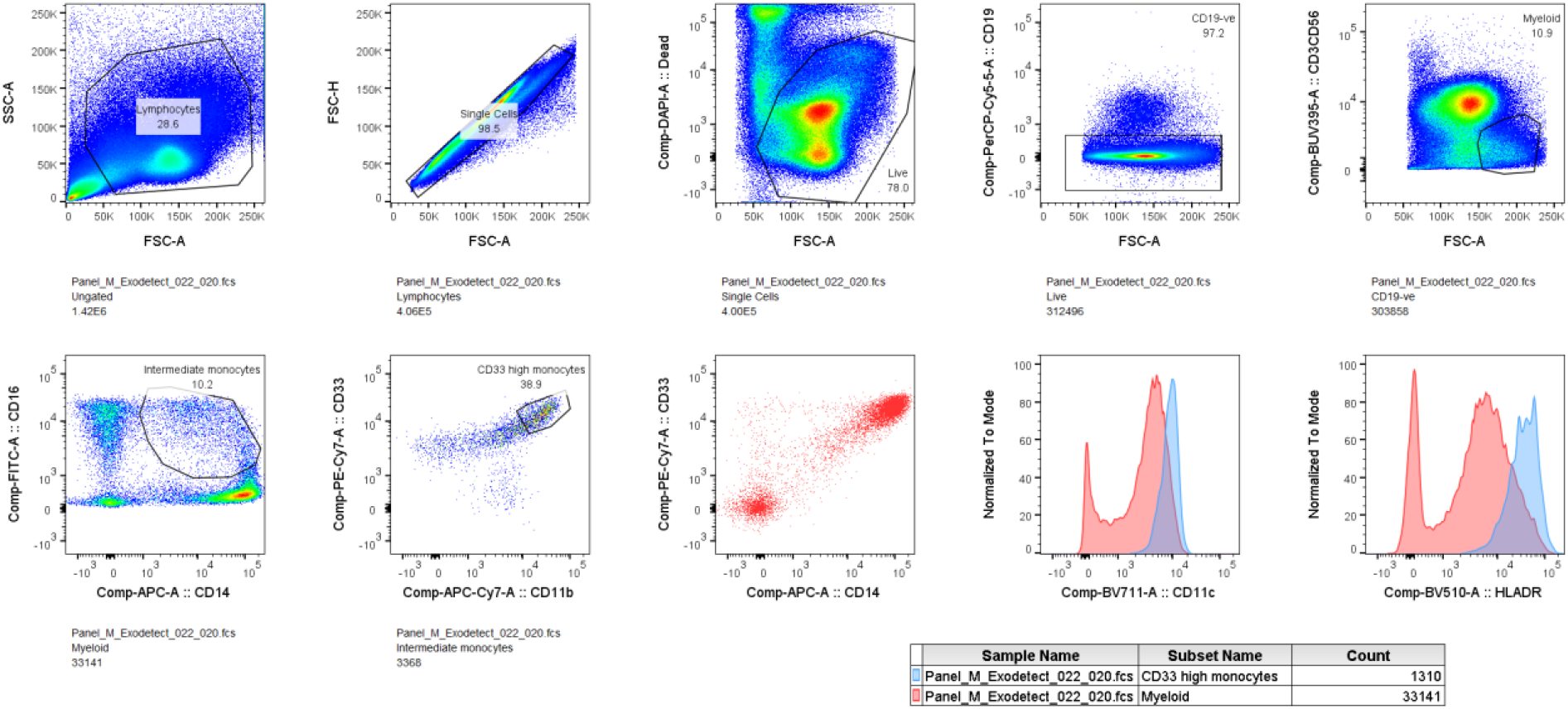
Gating strategy for further characterisation of CD33+CD14+ monocytic population using new patient cohort. CD14+CD16lowCD33+CD11b monocytes also express high levels of HLADR and CD11c.

**Supplementary Figure 4.**
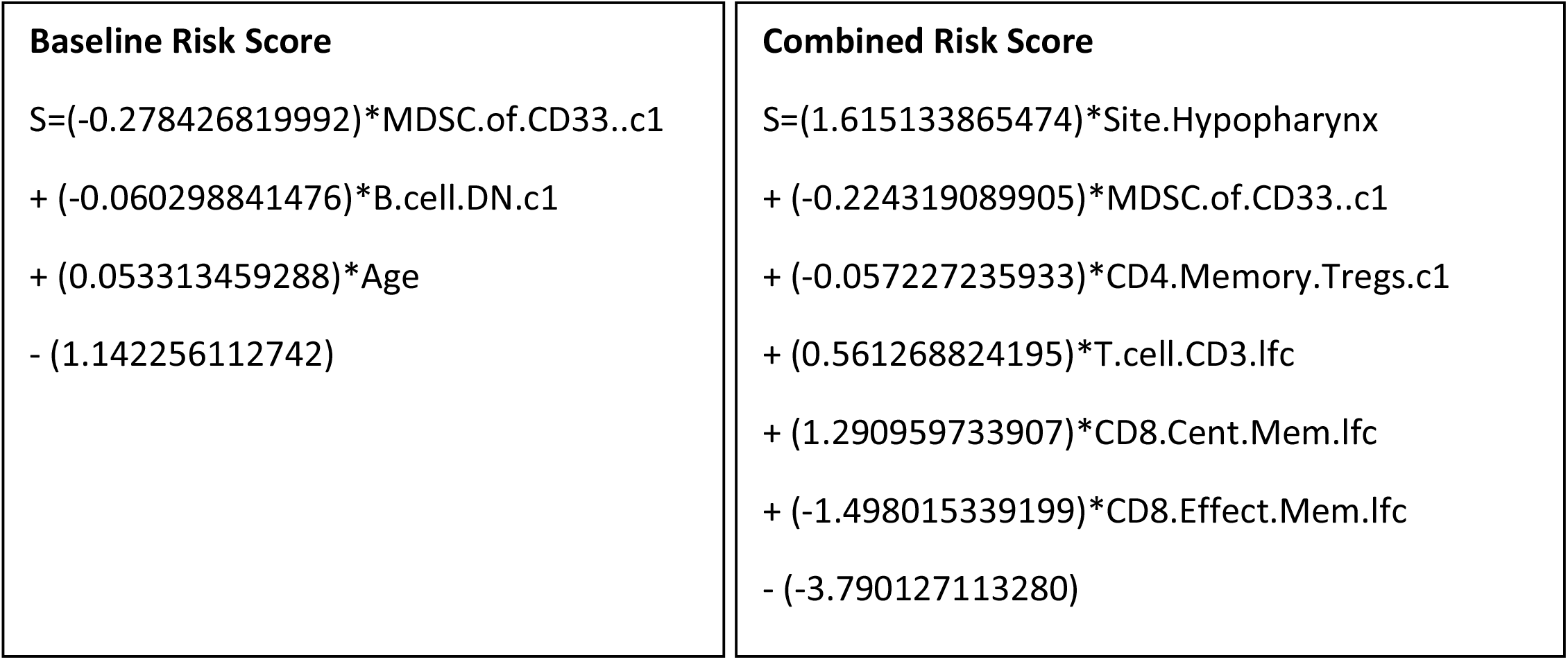
Risk score signatures for use with raw covariate values, with missing data imputed with the study mean from Supplementary Table 1. Use these equations to calculate the risk score for each patient.

## List of abbreviations

C1 or C2: Timepoint before cycle 1 or cycle 2 of treatment
C-index: Harrell’s Concordance Index
CT: Computed Tomography
CyTOF: Mass Cytometry
ddPCR: Digital Droplet Polymerase Chain Reaction
DN: Double-Negative
EGFR: Epidermal Growth Factor Receptor
FLIM: Fluorescence Lifetime Imaging Microscopy
FRET: Förster Resonance Energy Transfer
HPV: Human Papilloma Virus
HNSCC: Head and Neck Squamous Cell Cancer
HR: Hazard Ratio
KM: Kaplan-Meier
LFC: Log Fold Change
miRNA: Micro-RNA
PBMC: Peripheral Blood Mononuclear Cells
PCR: Polymerase Chain Reaction
PFS: Progression Free Survival
Tregs: Regulatory T cells

